# The Association Between Hypoglycemic Agents and Clinical Outcomes of COVID-19 in Patients with Diabetes: A Systematic Review and Meta-Analysis

**DOI:** 10.1101/2021.01.26.21250506

**Authors:** Tiantian Han, Shaodi Ma, Chenyu Sun, Huimei Zhang, Guangbo Qu, Yue Chen, Ce Cheng, Eric L. Chen, Mubashir Ayaz Ahmed, Keun Young Kim, Reveena Manem, Mengshi Chen, Zhichun Guo, Hongru Yang, Yue Yan, Qin Zhou

## Abstract

**Background:** During the current Coronavirus Disease 2019 (COVID-19) pandemic, diabetic patients face disproportionately more. Anti-inflammatory effects of hypoglycemic agents have been reported, and their beneficial or harmful effects in patients with diabetes and COVID-19 remain controversial.

**Purpose:** This study was performed to clarify this association.

**Data Sources:** Relevant literature was searched on China National Knowledge Infrastructure (CNKI), Wanfang Data Knowledge Service Platform, Chinese periodical service platform VIP Database, Sinomed (China Biology Medicine, CBM), MedRxiv, PubMed, ScienceDirect, Web of Science, Ovid Databases (LWW), Springer Link, Wiley Online Library, Oxford Academic, Nature Press Group, Cochrane Library and BMJ Evidence-Based Medicine up to November 14, 2020.

**Study Selection:** Only observational studies of hypoglycemic agents vs. drugs or therapy without hypoglycemic agents in adult diabetic patients with COVID-19 were included.

**Data Extraction:** Data of death and poor composite outcomes were extracted.

**Data Synthesis:** The pooled effects were calculated using the fixed-effects or random-effects models based on heterogeneity assessment.

**Limitation:** Most studies were retrospective cohort studies with relative weak capability to verify causality.

**Conclusion:** Home use of metformin might be beneficial in decreasing mortality in diabetic patients infected with SARS-CoV-2. There is insufficient evidence to conclude that metformin and other hypoglycemic agents are associated with poor composite outcomes. More prospective studies, especially RCTs are needed.

**Registration-PROSPERO:** CRD42020221951.

## 1. Introduction

During the current Coronavirus Disease 2019 (COVID-19) pandemic, there are a limited number of medications evidenced to be effective in treating COVID-19 patients(1). Although vaccines have been proved effective(2) and received emergent use authorization in some countries, their long-term protective effect is uncertain and unable to protect the already infected population(3-5). The global number of test-positive cases and deaths caused by severe acute respiratory syndrome coronavirus 2 (SARS-CoV-2) continues to increase. Accumulating evidence suggests that the release of a large amount of pro-inflammatory cytokines known as “cytokine storm” triggered by host immune response to the SARS-CoV-2 correlates directly with poor prognosis of COVID-19(6). Diabetes mellitus was reported as a major comorbidity, ranking after hypertension and cardiovascular disease(7). The prospective Dutch COVID-PREDICT cohort showed that the presence of hypertension, dyslipidemia and diabetes led to a stepwise increased risk for short-term mortality in hospitalized COVID-19 patients independent of age and sex(8), and similar results were reported by China CDC Weekly(9) and American CDC(10). A meta-analysis containing a total of 6,452 patients from 30 studies showed that diabetes was associated with increased mortality, severe illness, Acute Respiratory Distress Syndrome (ARDS), and disease progression in patients with COVID-19, and this association might be connected by inflammatory response(11).

Hypoglycemic agents including metformin, dipeptidyl-peptidase 4 inhibitors (DPP-4i), sulfonylurea, glinides, sodium-glucose co-transporter 2 inhibitors (SGLT-2i), glucagon-like peptide-1 receptor agonists (GLP-1RA), α-glycosidase inhibitors and thiazolidinediones (TZDs) have been approved, effective, generally safe, and widely used in treating diabetes(12, 13). Beyond their antidiabetic role, anti-inflammatory and antiviral effects of hypoglycemic agents have been noticed. Metformin, the most prescribed and first-line drug for diabetes, was evaluated as adjuvant therapy for patients with COVID-19(14). Five clinical trials have been registered thus far, and previous reports of effects of metformin on clinical prognosis of COVID-19 remain controversial(15, 16). DPP-4i, another commonly used antidiabetic drug, might not protect people from infection as reported in a large case-control study in which DPP-4i was more prevalent in diabetic patients with confirmed COVID-19 than those without COVID-19, but might still play an important role in protecting COVID-19 patients from organ failure and evolving pneumonia to pulmonary fibrosis(17). A clinical trial is ongoing in France to assess the efficacy of several repurposed drugs against COVID-19 including repaglinide(18). A propensity-score-matched cohort study also noted the influence of SGLT-2i on susceptibility to COVID-19 in diabetic patients(19). Unsolved queries remain about the effects of GLP-1RA, α-glycosidase inhibitors, and TZDs in patients with COVID-19. Thus we conducted this systemic review and meta-analysis to further explore queries about effects of hypoglycemic agents on mortality and poor composite outcomes of COVID-19 in diabetic patients.

## 2. Methods

A systematic review and meta-analysis was conducted under guidance of Meta-analysis Of Observational Studies in Epidemiology (MOOSE)(20). The PROSPERO registration number is CRD42020221951.

### 2.1 Data Sources and Searches

A systematic literature search was performed on China National Knowledge Infrastructure (CNKI), Wanfang Data Knowledge Service Platform, Chinese periodical service platform VIP Database, Sinomed (China Biology Medicine, CBM), MedRxiv, PubMed, ScienceDirect, Web of Science, Ovid Databases (LWW), Springer Link, Wiley Online Library, Oxford Academic, Nature Press Group, Cochrane Library, and BMJ Evidence-Based Medicine up to November 14, 2020. Search terms included hypoglycemic agents, metformin, DPP-4i, sulfonylurea, glinides, SGLT-2i, GLP-1RA, α-glycosidase inhibitors and TZDs and COVID-19. Exact search strategy was shown in Table S1. Duplicate results were removed automatically and manually. We screened the remaining articles by reading the title and abstract according to the inclusion and exclusion criteria. And disagreements were solved by discussion. Residual articles were assessed by full text.

### 2.2 Study Selection

Included studies needed to meet the following criteria: 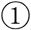observational study including cohort study, case control study and case series; 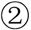patients were older than 18 years of age and diagnosed with both diabetes and COVID-19; 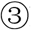home use or in-hospital use of specific hypoglycemic agents (including metformin, DPP-4i, sulfonylurea, glinides, SGLT-2i, GLP-1RA, α-glycosidase inhibitors and TZDs) vs. drugs or therapy except specific hypoglycemic agents; 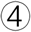 clinically validated definition of death, poor composite outcomes including intubation ventilation, Acute Respiratory Distress Syndrome (ARDS), Disseminated intravascular coagulation (DIC), intensive care unit (ICU) admission, disease progression or other adverse outcomes. The following articles were excluded: 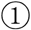 repeated research; 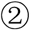 effective data is not applicable; 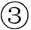could not obtain full text.

### 2.3 Data Extraction and Quality Assessment

Before collecting data, quality assessment was performed by using Newcastle-Ottawa Quality Assessment Scale(21). We developed a standardized extraction form to collect the following data: first author’s name, year, study date, study geographical region, study design, sample size, male (percentage), groups, death number or unadjusted OR of mortality, adjusted OR/HR of mortality, adjustment factors, poor composite outcomes, data source, home/in-hospital use drugs and NOS stars. Hazard ratios (HRs) were broadly equivalent to risk ratios (RRs) when the risk is not constant with respect to time and RRs were transformed to ORs before merging(22, 23). Data was extracted and checked. Disagreements were resolved by discussion.

The outcomes of interest were clinically validated definition of death, poor composite outcomes comprised of intubation ventilation, Acute Respiratory Distress Syndrome (ARDS), Disseminated intravascular coagulation (DIC), intensive care unit (ICU) admission, disease progression or other adverse outcomes. We contacted one author for further information that was not presented in detail in one of the published studies.

### 2.4 Data Synthesis and Analysis

Review Manager 5.3 (Cochrane Collaboration) and Stata 14.0 (Stata, version 14; Stata Corp, College Station, TX, USA) were used for meta-analysis. The effect estimate was reported as pooled odd ratios (ORs) along with its 95% confidence intervals (95% CIs) by generic inverse variance. To reflect the variation in treatment effects over different settings and the expected effect in future patients, the prediction intervals (PIs) were reported as a supplement to CIs(24). Pooled ORs, CIs and PIs of mortality, and poor composite outcomes were calculated. Two-tailed P≤0.05 was considered statistically significant. Q test and I^2^ statistic was performed to judge heterogeneity. I^2^ < 50% and P>0.1 indicated low statistical heterogeneity, and a fixed effects model would be used; I^2^ ≥ 50% and P<0.1 indicated exact heterogeneity, and a random effects model would be used. 95% CI of I^2^ was reported for further heterogeneity assessment. Funnel plot and Egger’s test was done to investigate for publication bias. Sensitivity analysis was performed to explore the source of heterogeneity and determine subsequent analysis. If applicable, subgroup analysis was performed between adjusted or unadjusted ORs, different regions, home or in-hospital use of hypoglycemic agents, drug vs. non-drug in patients with type 2 diabetes only, respectively.

## 3. Results

### 3.1 Study selection and characteristics

A total of 925 records were identified through database searching, and 681 records remained after duplicates were removed. After screening the titles and abstracts, 47 records remained and were evaluated by full text. Afterwards, 31 records were excluded for the following reasons: 7 articles were unrelated; 12 articles related to mechanism; 8 articles had no clear comparable groups; 3 articles had no related outcomes; 2 articles were repetitive. Finally, the remaining 16 studies were included for qualitative synthesis, and 14 studies with 13,371 patients were included in quantitative synthesis (meta-analysis). The flow chart is shown in Figure 1.

**Figure 1.**
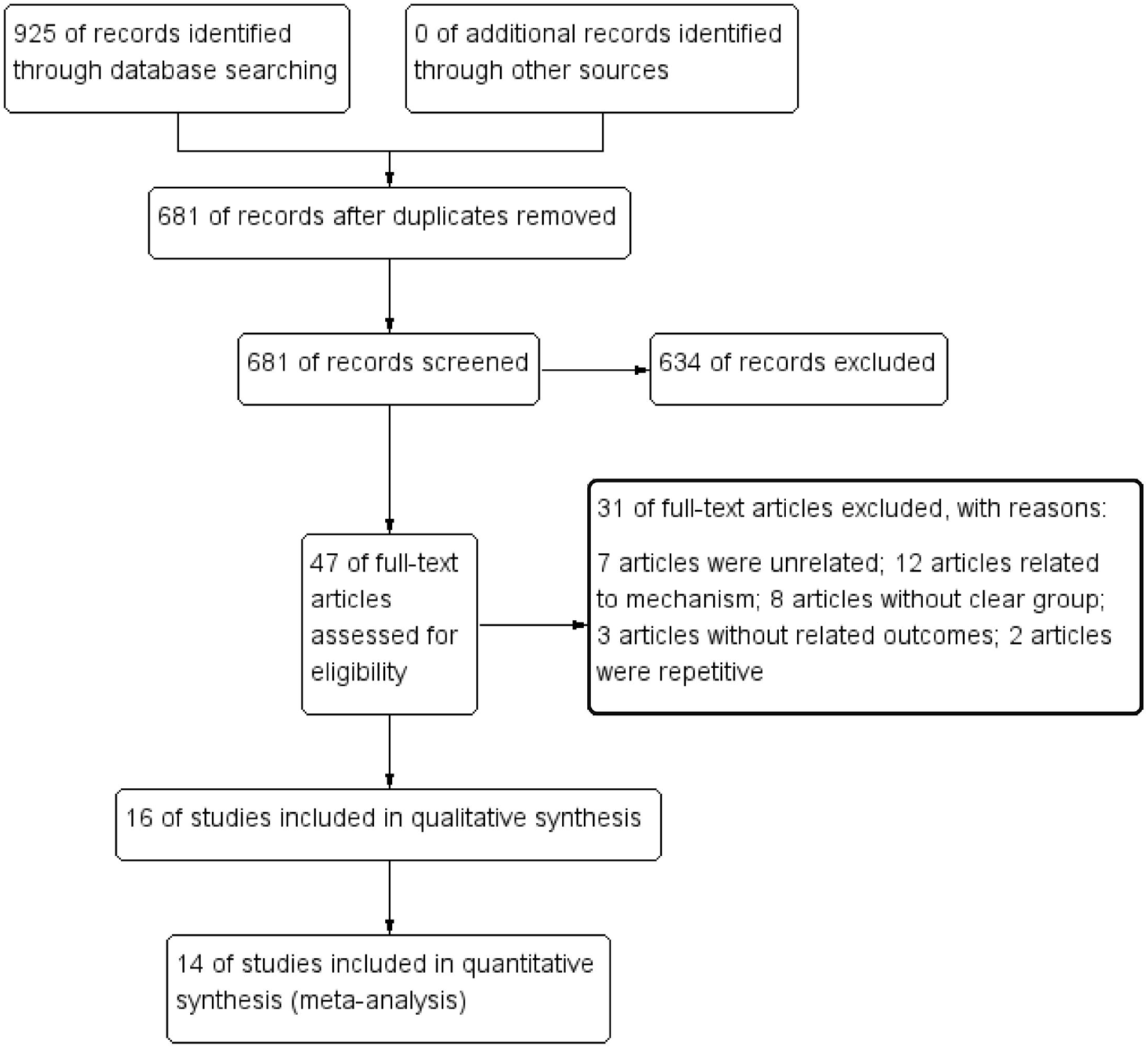
Flow diagram of study selection processes.

16 studies were included in qualitative synthesis. Among these, 12 studies related to metformin(15, 16, 25-35), 6 studies related to DPP-4i(27, 28, 33, 35-38), 3 studies related to sulfonylurea or glinides(27, 28, 33, 35), 2 studies related to SGLT-2i(33, 35) and 1 study related to GLP-1RA(27, 28). Some of these studies included multiple outcomes to different medications. 14 studies were included for quantitative synthesis, among which, 11 studies investigated metformin, and 5 studies investigated DPP-4i. All included studies were assessed with ≥6 stars by NOS. All studies included in quantitative synthesis were cohort studies, and most were electronic medical records collected by clinicians with high authenticity and credibility. Details are shown in Table 1.

**Table 1.**
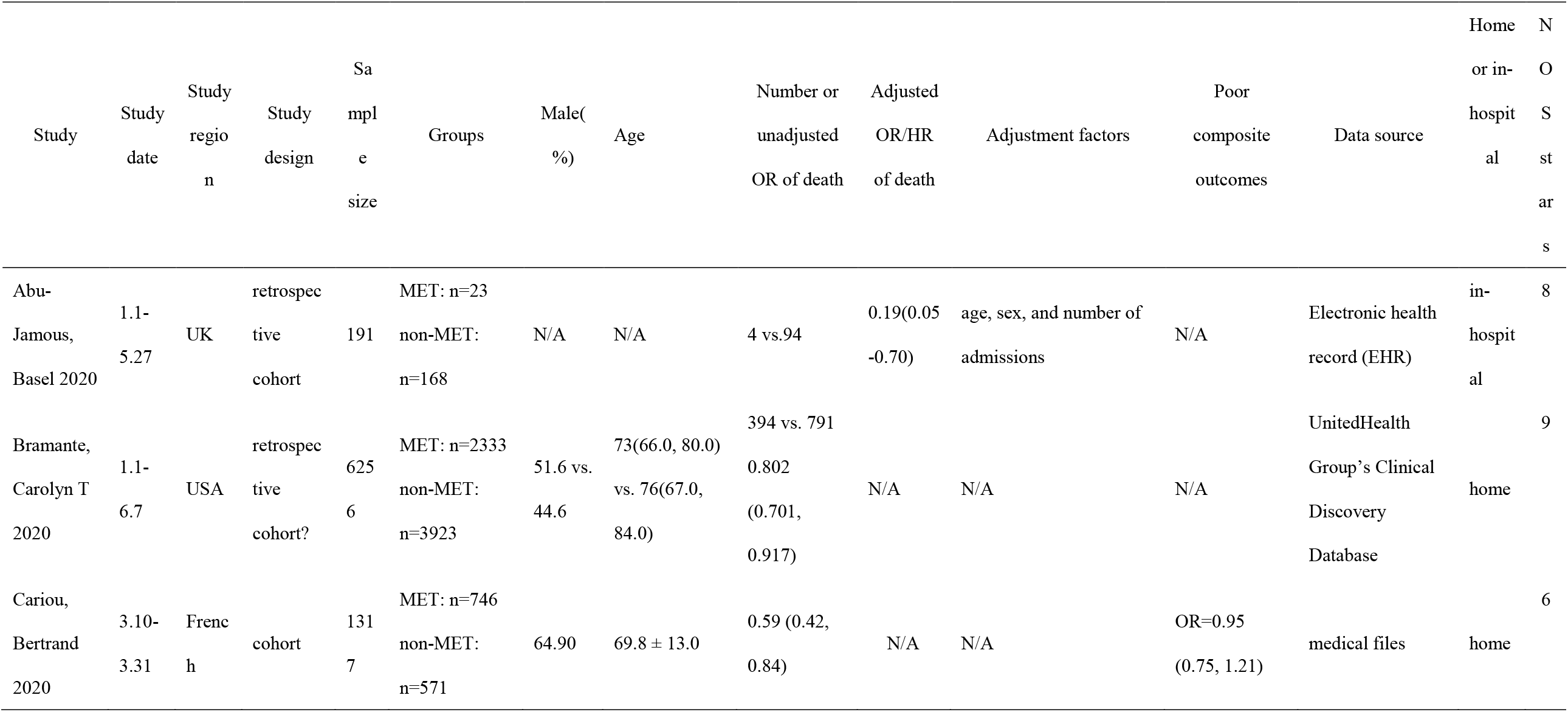
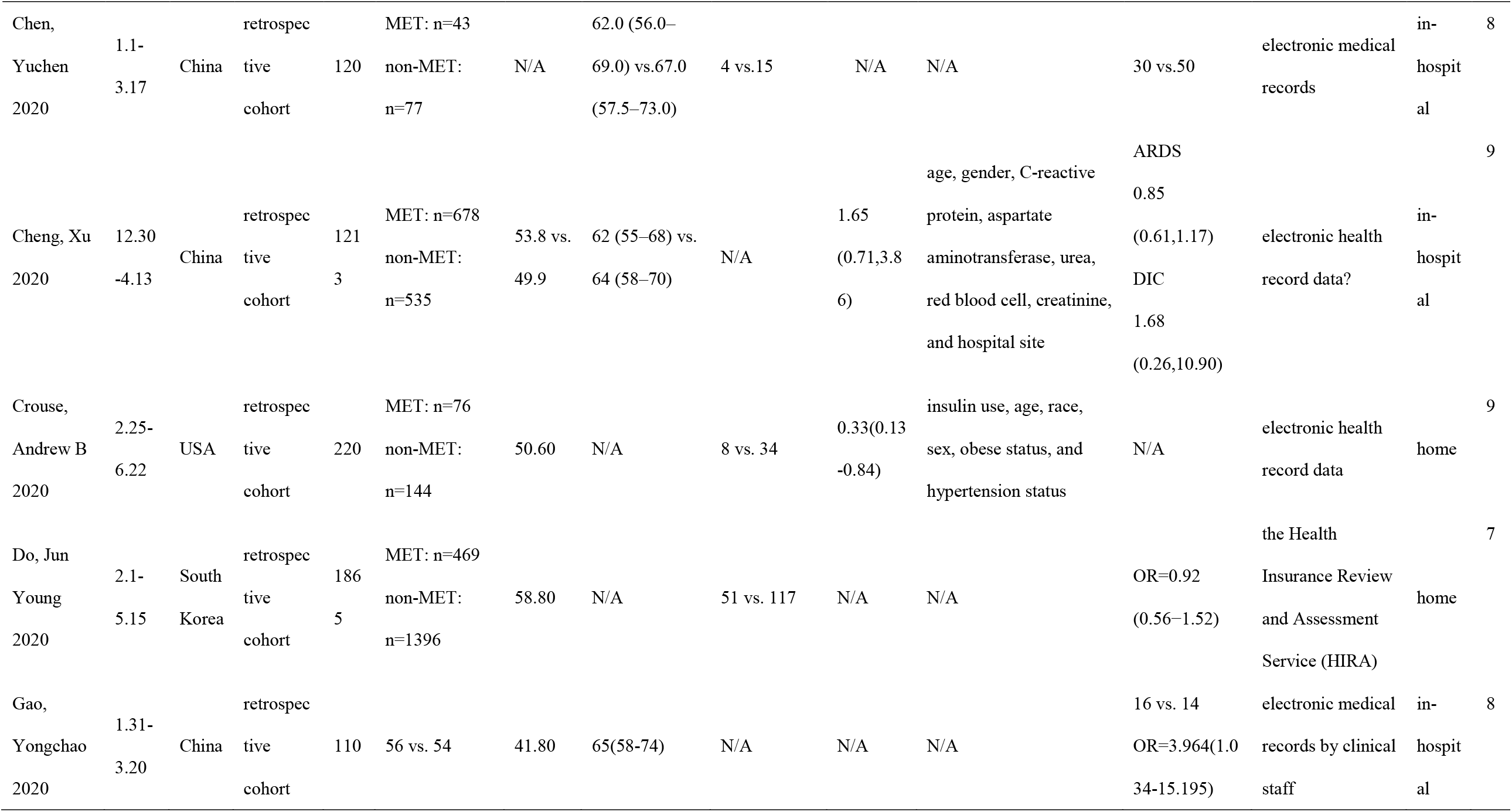
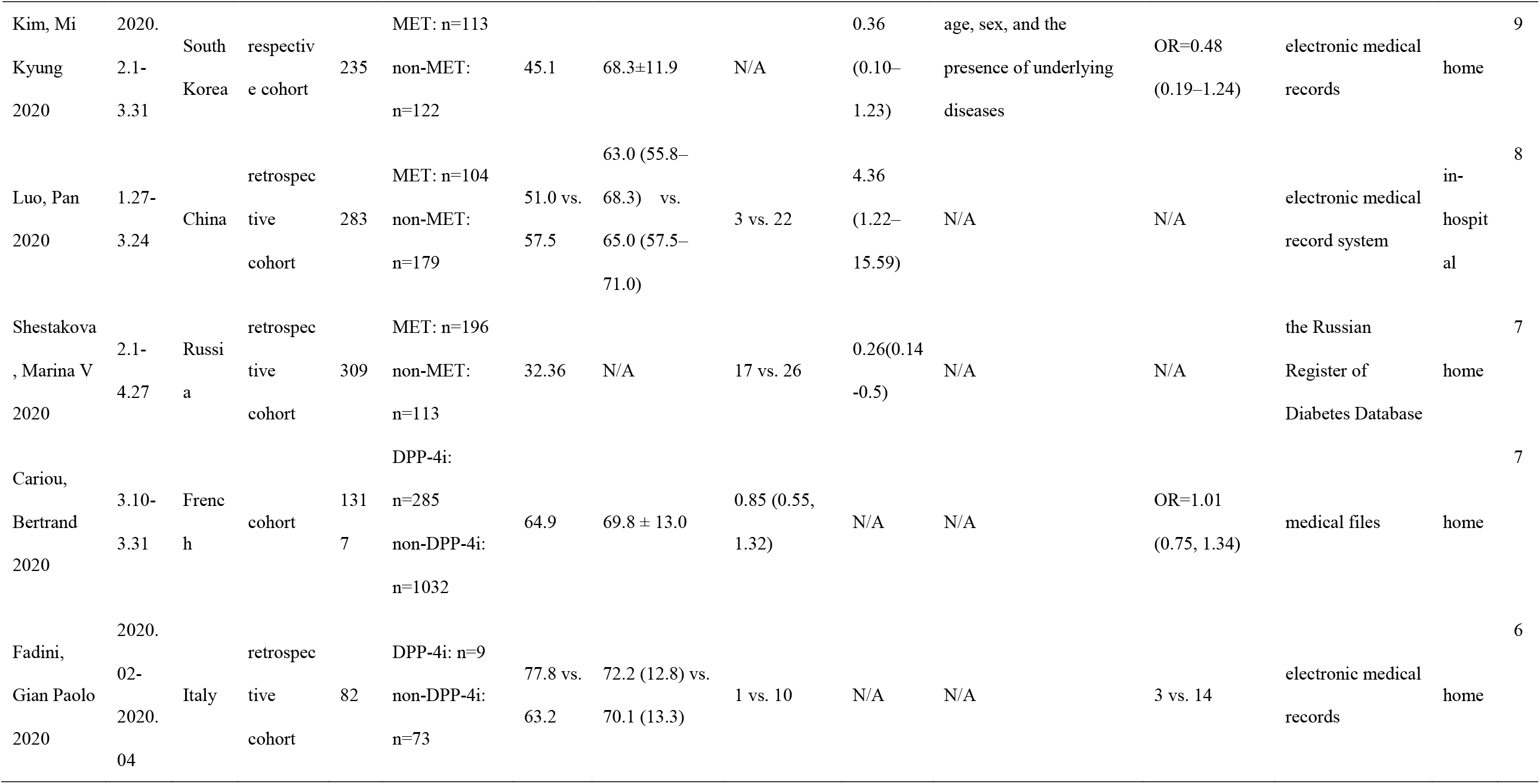
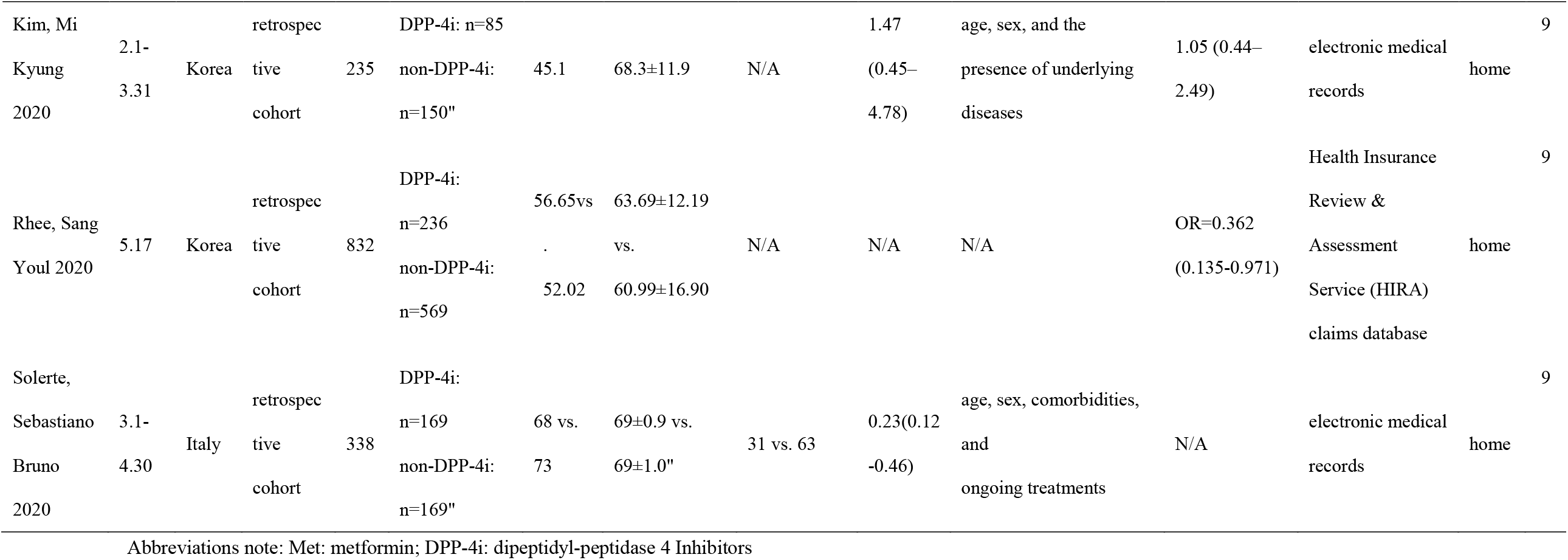
The main characteristics of studies included in the meta-analysis

### 3.2 Effects of metformin vs. non-metformin on mortality and poor composite outcomes of COVID-19 in diabetic patients

#### 3.2.1 Effects of metformin vs. non-metformin on mortality

Meta-analysis of 10 cohort studies investigating mortality risk of COVID-19 for diabetic patients taking metformin showed that metformin was associated with a statistically significant reduced mortality risk (pooled OR=0.56, 95% CI, 0.39-0.81, P=0.002, I^2^=77%, P<0.0001, 95% CI of I^2^, 57%-83%, 95% PI, 0.56-4.79) (Figure 2).

**Figure 2.**
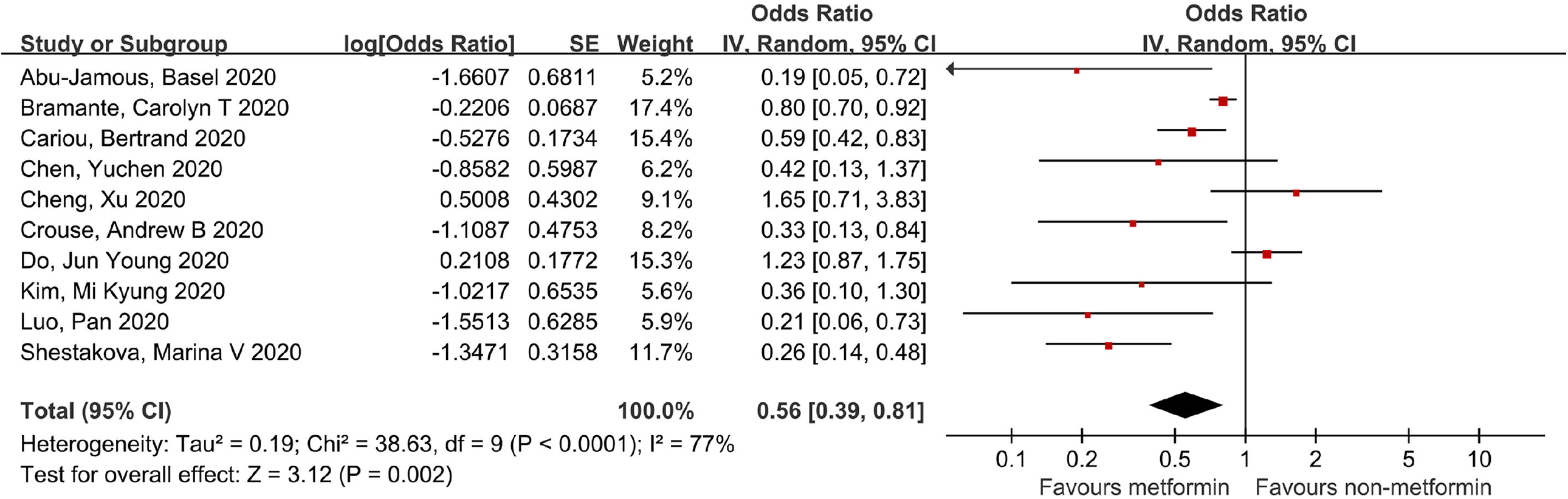
Forest plot of the association between metformin and mortality risk of COVID-19 for diabetic patients.

#### 3.2.2 Subgroup analysis of effect of metformin vs. non-metformin on mortality

Subgroup analysis was performed between adjusted and unadjusted data, home use and in-hospital use of medication, different geographical regions, and metformin vs. non-metformin in patients with type 2 diabetes only to explore potential confounders and expose biases.

After adjustment by age, gender, comorbidities, etc., metformin use was still associated with a statistically significant reduced mortality risk in diabetic patients with COVID-19 (pooled adjusted OR=0.41, 95% CI, 0.19-0.91, P=0.03, I^2^=72%, P=0.007, 95% CI of I^2^, 28%-89%, 95% PI, 0.18-14.74) (Figure S1).

Home use of metformin was associated with a statistically significant reduced death risk (pooled OR=0.60, 95% CI, 0.40-0.88, P=0.01, I^2^=81%, P<0.0001, 95% CI of I^2^, 59%-91%, 95% PI, 0.52-5.19), but in-hospital use of metformin did not show such an association (pooled OR=0.44, 95% CI, 0.15-1.33, P=0.14, I^2^=74%, P=0.01, 95% CI of I^2^, 26%-91%, 95% PI, 0.05-55.90) (Figure 3).

**Figure 3.**
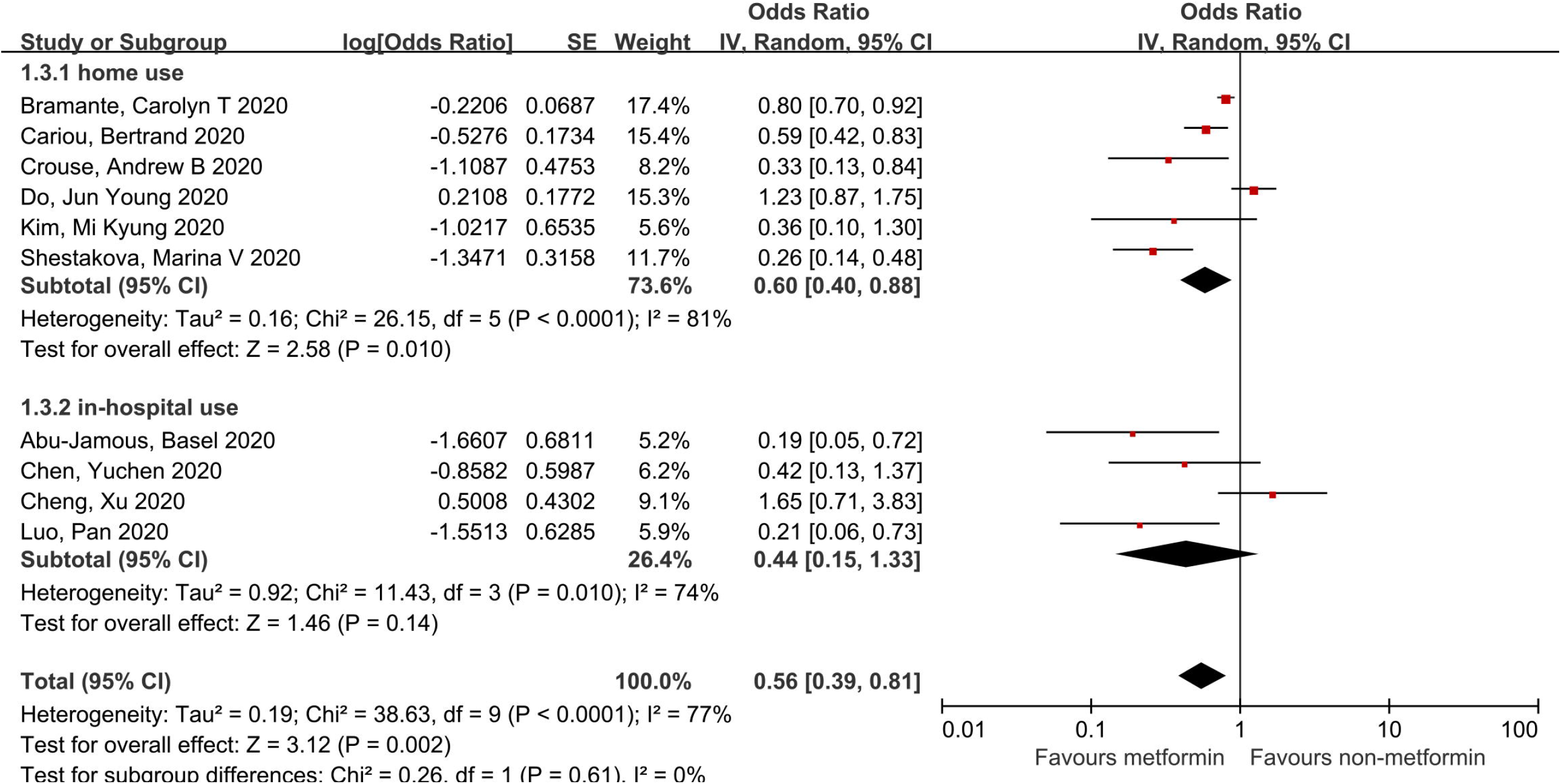
Forest plot of the association between metformin and mortality risk of COVID-19 for diabetic patients: home vs. in-hospital use of medication.

The subgroup analysis based on the study locations showed a statistically significant reduced mortality risk of COVID-19 among diabetic patients using metformin in Europe (pooled OR=0.36, 95% CI, 0.17-0.73, P=0.005, I^2^=71%, P=0.03, 95% CI of I^2^, 3%-92%, 95% PI, 0.11-25.63), but not in Asia (pooled OR=0.68, 95% CI, 0.33-1.40, P=0.29, I^2^=70%, P<0.01, 95% CI of I^2^, 24%-88%, 95% PI, 0.20-13.48) or North America (pooled OR=0.58, 95% CI, 0.25-1.34, P=0.21, I^2^=71%, P<0.06, 95% CI of I^2^, -30%-93%, 95% PI, 0.09-30.69) (Figure S2).

A subgroup analysis of metformin vs. non-metformin in patients with only patients with type 2 diabetes and COVID-19 showed that metformin was not associated with a statistically significant reduced death rate (pooled OR=0.82, 95% CI, 0.52-1.29, P=0.39, I^2^=78%, P=0.01, 95% CI of I^2^, 28%-93%, 95% PI, 0.29-9.35) (Figure S3).

#### 3.2.3 Publication bias assessment

A funnel plot was drawn to judge the publication bias (Figure S3). Egger’s test was performed to quantitatively assess the publication bias (t=-1.78, p=0.112>0.05). The results showed no publication bias (Figure S5).

#### 3.2.4 Sensitivity analysis

We detected significant heterogeneity in analysis of the association between metformin and mortality risk of COVID-19 for diabetic patients (Q test, P<0.0001, I^2^=77%, 95% CI of I^2^, 57%-87%). To identify the influence of individual studies on combined effects, sensitivity analysis was performed by excluding each original study. The fluctuation of the pooled ORs was found to be between 0.30 and 0.91, with lower limit and upper limit of 95% CI constantly less than 1, and P-value constantly less than 0.05, suggesting the stability of this meta-analysis. (Figure S6).

#### 3.2.5 Effects of metformin vs. non-metformin on poor composite outcomes

Considering different studies reported different outcomes(15) (27, 28) (29) (31) (32) (33) (Table S2), we defined poor composite outcomes to include intubation ventilation, ARDS, DIC, ICU admission, disease progression, and other adverse outcomes (31, 32). Studies reporting any of these outcomes were included in combined analysis. No statistical significance was found between metformin and poor composite outcomes (pooled OR=0.93, 95% CI, 0.79-1.11, P=0.44, I^2^=21%, P=0.27) (Figure S7)

### 3.3 Effects of DPP-4i vs. non-DPP-4i on mortality and poor composite outcomes of COVID-19 in diabetic patients

DPP-4i were associated with statistically non-significant reduced risk of mortality (pooled OR=0.63, 95% CI, 0.26-1.56, P=0.32, I^2^=77%, P=0.005, 95% CI of I^2^, 37%-92%, 95% PI, 0.10-27.19) (Figure S8) and poor composite outcomes of COVID-19 in diabetic patients (pooled OR=0.96, 95% CI, 0.74-1.26, P=0.78, I^2^=39%, P=0.17) (Figure S9).

### 3.4 Effects of other hypoglycemic agents on prognosis of COVID-19 in diabetic patients

Qualitative analysis was conducted to evaluate other hypoglycemic agents including sulfonylurea, glinides, SGLT-2i, GLP-1RA, α-glycosidase inhibitors and TZDs. Quantitative analysis was not performed due to insufficient data. Three studies investigated sulfonylurea’s effects on death. One study investigated the combined use of sulfonylurea and glinides, and found no statistically significant effect on death or tracheal intubation(27). In addition, no statistically significant result of sulfonylurea’s effect on death or severe disease was found in another study(33), and the last was a case-series study without reporting group outcomes(35). Eight patients in Kim’s study and one patient in Xu’s study received SGLT-2i, with the former reporting no statistically significant effect on death or severe disease(33) and the later didn’t report result(35). The results may have been limited by the small sample size. Cariou’s study also included patients who take GLP-1RA; however, no statistically significant effect on death or tracheal intubation was found. We did not find original studies related to α-glycosidase inhibitors or TZDs.

## 4. Discussion

As reported in a retrospective cohort of hospitalized patients in the UK, long-term antidiabetic medications reduced COVID-19 mortality in diabetic patients(25), which was a very encouraging finding, especially as diabetic patients are particularly susceptible to cumulative organ injury by SARS-CoV-2 because of already compromised pulmonary, cardiac and renal functions. Therefore, furthering our understanding of antidiabetic medications can yield a practical and effective approach in dramatically improving outcomes in this vulnerable population disproportionately affected by COVID-19.

The result of our meta-analysis showed that metformin was associated with a statistically significant reduced mortality for diabetic patients with COVID-19, both before and after adjusting for other factors. This was consistent with several previous small-scale meta-analyses(39, 40). Home use of metformin was more effective compared to in-hospital use, which was shown in previous analyses(41). This might be explained by side effects, e.g. an increased risk for metabolic acidosis(15), induced by metformin(42) in hospitalized patients. In patients with organ function compromised by COVID-19, the benefits of metformin were likely to be overshadowed by its side effects(15, 43). As a result, metformin was discontinued and switched to insulin for inpatients in many countries. Benefits of metformin on mortality were more obvious in Europe compared to Asia or North America according to our analysis, although the benefits of metformin were still evident in the United States. For example, a large retrospective review from UnitedHealth Group’s Clinical Discovery Database in the USA reported that home use of metformin was significantly associated with reduced mortality in female inpatients with COVID-19, and reduction of TNF-α might underly the mechanistic pathway(26). In addition, confounders such as race, gender, and disease severity require further study. Metformin had no beneficial effects on poor composite outcomes including tracheal intubation, ARDS, DIC, ICU admission, and disease progression, indicating that metformin might be more effective in reducing mortality risk in severe or critical illness, while not preventing disease progression to severe or critical illness. Current discussions on metformin’s role in reducing COVID-19 mortality involves several different mechanisms. One possible mechanism is that metformin inhibits cytokine storm by suppressing interleukin-6 (IL-6) signaling, preventing the process of lung fibrosis and suppressing endocytosis, thereby elevating angiotensin converting enzyme 2 (ACE2) expression(44). Increased ACE2 offers cardiopulmonary benefits in patients with COVID-19 by activating AMP-activated protein kinase (AMPK), which is involved in phosphorylating key molecules regulating metabolism and cardiovascular health(45). Activated AMPK phosphorylates ACE2, thereby changing conformation and function of ACE2 and resulting in decreased integration with the SARS-CoV-2 receptor and binding domain due to steric hindrance(46). The mortality benefits may also be explained by metformin’s effect on cellular pH, which needs to be acidic for optimal viral membrane fusion of SARS-CoV-2. By increasing cellular pH, metformin subsequently interferes with the endocytic cycle to inhibit viral infection by acting on the eNHEs and/or the V-ATPase(14). Reducing production of proinflammatory cytokines by macrophages and formation of neutrophil extracellular traps (NETs) might play a role as well(43). Although current evidence supports metformin use, further studies are needed to understand its mechanistic link to mortality benefits.

Use of DPP-4i for diabetic patients is not associated with statistically non-significant reduced death or poor composite outcomes of COVID-19 according to our analysis. Previous studies comparing the prevalence of DPP-4i use between diabetic patients with or without COVID-19 showed that DPP-4i use was more prevalent in COVID-19 patients(47), implying that DPP-4i might not prevent people from contracting SARS-CoV-2(48). However, some opinions indicated that diabetic patients with COVID-19 could benefit from DPP-4i not only by controlling blood glucose, but also improving long-term prognosis of COVID-19 caused by pulmonary fibrosis, heart and kidney injury via blocking the tissue remodeling function of activating myofibroblasts and migrating fibroblasts, suppressing inflammatory sign and proliferating vascular smooth muscle cells to avoid adverse outcomes(17). As DPP-4 is hypothesized to be a binding partner for corona-like viruses to enter host immune cells, DPP-4i also exerts influence on prohibiting invasion of SARS-CoV-2 into cells(49). However, concentration of circulating soluble DPP-4 serum in patients suffering from severe COVID-19 was significantly lower compared to that in healthy human subjects(50), which contradicts the protective effects of DPP-4i. Sitagliptin, a DPP-4i drug, was thought to have anti-inflammatory effects on diabetic patients via the NF-kappa-B signaling pathway (51). This hypothesis needs to be verified by more studies.

Existing studies of other hypoglycemic agents, including sulfonylurea, glinides, SGLT-2i, GLP-1RA, α-glycosidase inhibitors, and TZDs, were insufficient for quantitative analysis. Apart from existing clinical studies, an *in vitro* experiment found that E protein, a potential ion channel on SARS-CoV-2, could be inhibited by Gliclazide (a type of sulfonylurea)(52). A propensity-score-matched cohort study showed that SGLT-2i did not decrease susceptibility compared to DPP-4i(19). Dapagliflozin, a type of SGLT-2i, was assumed to reduce the viral load by lowering of cytosolic pH(53). GLP-1RA was considered for treating asymptomatic and non-critically ill COVID-19 patients due to its anti-inflammatory effects(54) and this effect might be related with ACE2 as well. Pioglitazone, a common type of TZDs, was recommended to treat COVID-19 patients for its potential to improve liver injuries(55) and block macrophage activation by uptake of oxidized LDL to reduce the progression of atherosclerosis, which results in less risk of developing into severe illness of COVID-19(56).

Heterogeneity existed in analysis of effects of metformin on mortality risk and effects of DPP-4i on mortality risk and poor composite outcomes. Even after subgroup analysis of adjusted and unadjusted data, home use and in-hospital use of medication, different geographic regions, and drug vs. non-drug in patients with type 2 diabetes only, heterogeneity was not eliminated or significantly reduced. Sensitivity analysis did not identify any specific original study that led to unstable results, which might be explained by the discrepancy of included studies themselves. Gender ratio and average age accounted for a portion of unstable results as female and young patients(30, 34) reportedly have a better prognosis. Elements including duration of medication (from <21 day(25) to >6 months), dose, common drugs in exposure and control groups(29), long-term blood glucose control (HbA1c)(34, 57), BMI(26) and comorbidities(58) varied in different studies and were likely related to heterogeneity. We also calculated the prediction interval (PI) to reflect the variation in treatment effects over different settings, predict the expected effect on future patients as a supplement to CI(24) and 95% CI of I^2^ described the proportion of total variation in study estimates that is due to heterogeneity (59). Results showed unstable predictive effects and heterogeneity which urges some caution about the findings.

In addition, there are several other potential limitations. Firstly, meta-analyses are inherently subject to design biases and variations of included original studies with discrepancies in methods and patient characteristics; despite this, statistically significant results were achieved by inclusion of large-sample studies and more original studies, as well as various dimensions and parameters of analysis to expose confounding variables and narrow this discrepancy. Although most studies contained in our analysis were electronic medical records collected by clinicians with high authenticity and credibility, most studies were retrospective cohort studies with relative weak capability to verify causality. As participants of Bramante’ study included both diabetic patients and patients with obesity(26), confounding factors such as obesity may have influenced the result. Participants in Cariou’s study were observed for only 7 days to draw conclusions, while longer follow-up period was preferred. We contacted the author for raw data, but as there was no response, the HR was directly merged with other studies’ OR which may marginally affect the pooled OR, but would not change the final conclusion.

## 5. Conclusion

Metformin is associated with reduced mortality of COVID-19 in diabetic patients, and no evidence of decreasing poor composite outcomes (intubation ventilation, ARDS, ICU admission, disease progression, and other adverse outcomes) has yet been found. Long-term home use of metformin is strongly supported because of its anti-inflammatory and antiviral effects. DPP-4i is not associated with a reduced risk of death or poor composite outcomes in diabetic patients with COVID-19. Current evidence is insufficient to draw a solid conclusion regarding the effects of sulfonylurea, glinides, SGLT-2i, GLP-1RA, α-glycosidase inhibitors or TZD on clinical outcomes of COVID-19 in diabetic patients. More prospective research studies, especially RCTs, are needed to verify and explore the effects of hypoglycemic agents on COVID-19 in diabetic patients.

## Data Availability

The raw data required to reproduce these findings cannot be shared at this time as the data also forms part of an ongoing study.

## 6. Acknowledgments

Author contributions: Tiantian Han and Dr. Chenyu Sun designed research; Tiantian Han, Shaodi Ma, Huimei Zhang and Guangbo Qu conducted literature search and data extraction; Tiantian Han, Shaodi Ma, Huimei Zhang, Guangbo Qu and Dr. Chenyu Sun analyzed data; Tiantian Han and Dr. Chenyu Sun wrote the paper. Dr. Chenyu Sun, Yue Chen, Dr. Ce Cheng, Dr. Mubashir Ayaz Ahmed, Keun Young Kim, Dr. Eric Chen, Dr. Reveena Manem, Mengshi Chen, Zhichun Guo, Hongru Yang, Yue Yan; Qin Zhou revised the paper. All authors read and approved the final manuscript. We appreciate all authors for their contributions, and support from AMITA Health Saint Joseph Hospital Chicago and Anhui Medical University.

Tiantian Han and Shaodi Ma had primary responsibility for final content. Dr. Chenyu Sun is the corresponding author. This study is financially supported by Hunan Provincial Key Laboratory of Clinical Epidemiology (grant number: 2020ZNDXLCL002). No conflict of interest.

## Notes

### Competing Interest Statement

The authors have declared no competing interest.

### Author Declarations

Registration-PROSPERO: CRD42020221951

