## Supplementary material for "The Association Between Hypoglycemic Agents and Clinical Outcomes of COVID-19 in Patients with Diabetes: A Systematic Review and Meta-Analysis": Figure S1, Figure S2, Figure S3, Figure S4, Figure S5, Figure S6, Figure S7, Figure S8, Figure S9, Table S1, Table S2

Background: During the current Coronavirus Disease 2019 (COVID-19) pandemic, diabetic patients face disproportionately more. Anti-inflammatory effects of hypoglycemic agents have been reported, and their beneficial or harmful effects in patients with diabetes and COVID-19 remain controversial.

Purpose: This study was performed to clarify this association.

Data Sources: Relevant literature was searched on China National Knowledge Infrastructure (CNKI), Wanfang Data Knowledge Service Platform, Chinese periodical service platform VIP Database, Sinomed (China Biology Medicine, CBM), MedRxiv, PubMed, ScienceDirect, Web of Science, Ovid Databases (LWW), Springer Link, Wiley Online Library, Oxford Academic, Nature Press Group, Cochrane Library and BMJ Evidence-Based Medicine up to November 14, 2020.

Study Selection: Only observational studies of hypoglycemic agents vs. drugs or therapy without hypoglycemic agents in adult diabetic patients with COVID-19 were included.

Data Extraction: Data of death and poor composite outcomes were extracted.

Data Synthesis: The pooled effects were calculated using the fixed-effects or random-effects models based on heterogeneity assessment.

Limitation: Most studies were retrospective cohort studies with relative weak capability to verify causality.

Conclusion: Home use of metformin might be beneficial in decreasing mortality in diabetic patients infected with SARS-CoV-2. There is insufficient evidence to conclude that metformin and other hypoglycemic agents are associated with poor composite outcomes. More prospective studies, especially RCTs are needed.

Registration-PROSPERO: CRD42020221951.

Keyword: Hypoglycemic Agents, Metformin, DPP-4 Inhibitors, Sulfonylurea, Glinides, SGLT-2 Inhibitors, GLP-1 Receptor Agonists, α-Glycosidase Inhibitors, Thiazolidinediones, COVID-19, SARS-CoV-2.

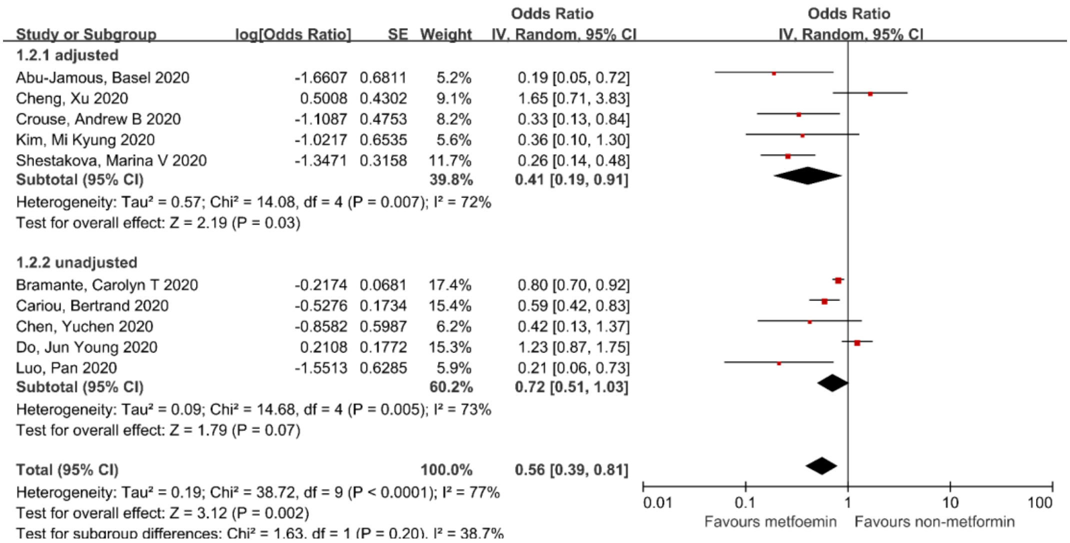


Figure S1. Forest plot of the association between metformin and mortality of COVID-19 for diabetic patients: adjusted vs. unadjusted data

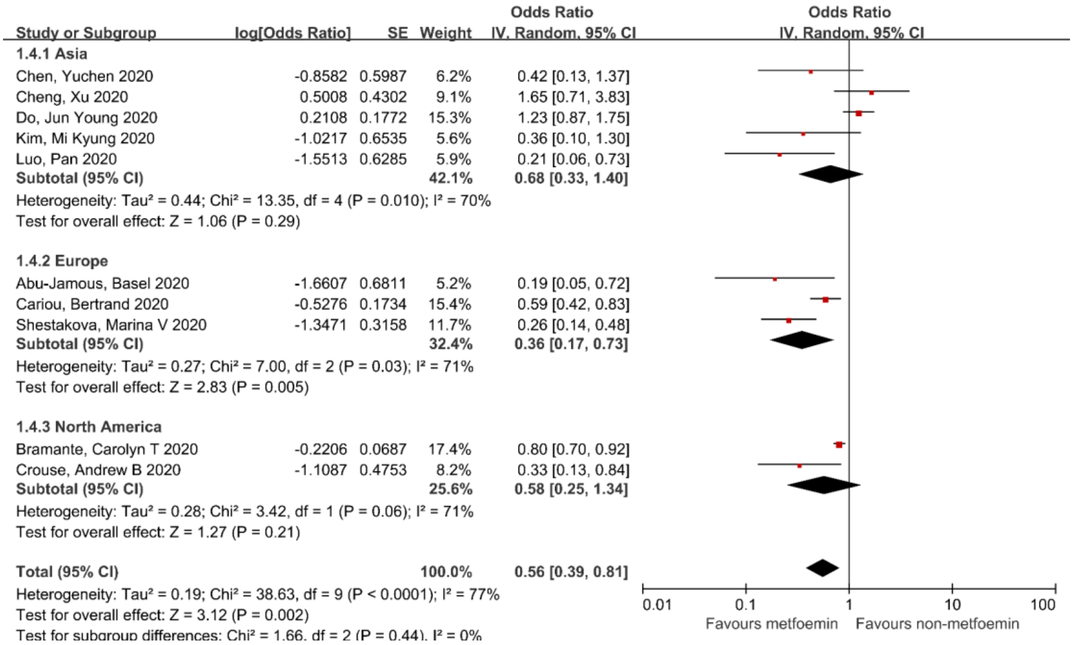


Figure S2. Forest plot of the association between metformin and mortality of COVID-19 for diabetic patients: different geographical regions

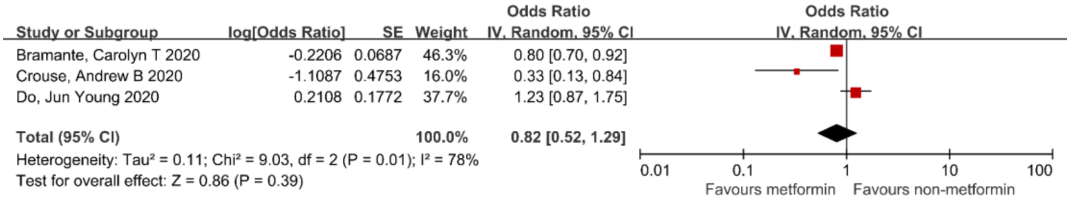


Figure S3. Forest plot of the association between metformin and mortality risk of COVID-19 for diabetic patients: metformin vs. non-metformin in patients with only patients with type 2 diabetes

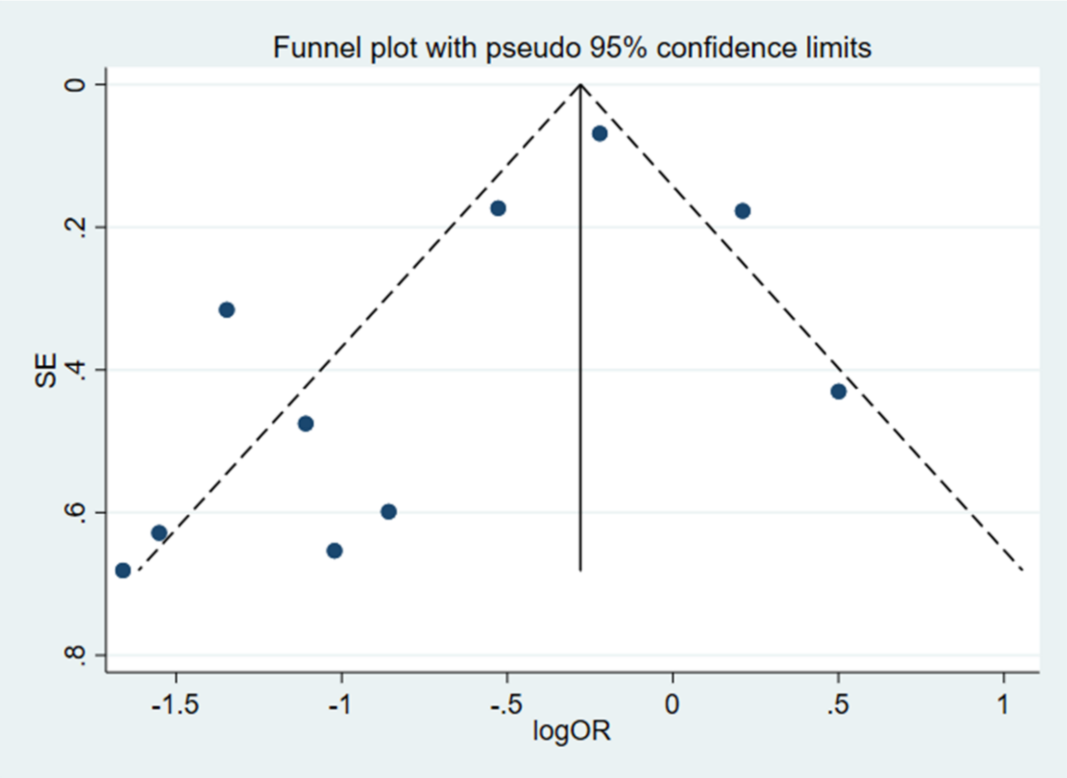


Figure S4. funnel plot of the association between metformin and mortality risk of COVID-19

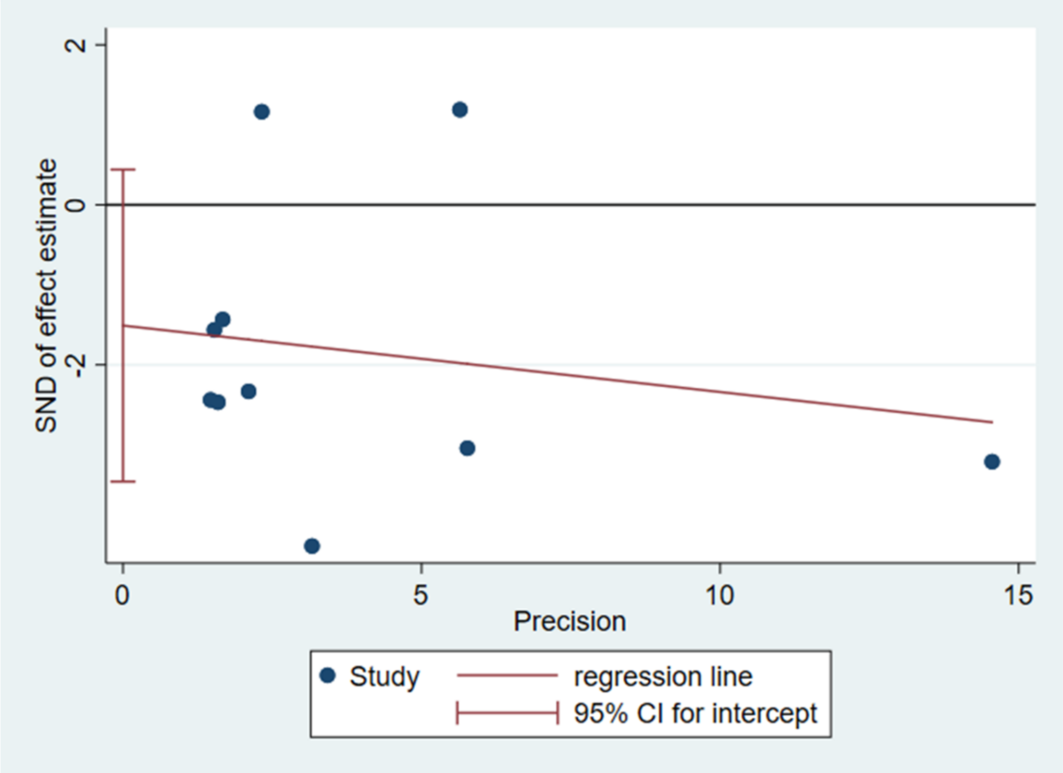


Figure S5. Egger's test of the association between metformin and mortality risk of COVID-19

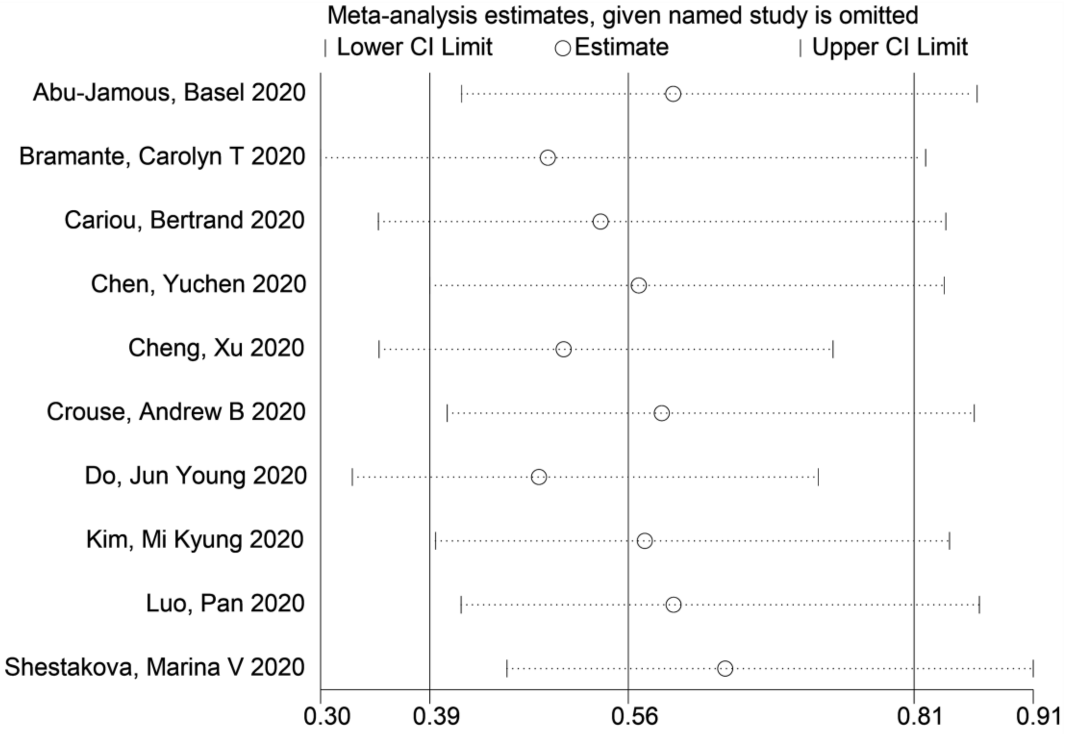


Figure S6. sensitivity analysis of the association between metformin and mortality risk of

| Study or Subgroup | log[Odds Ratio] | SE | Weight | Odds Ratio IV, Fixed, 95% CI |
|---|---|---|---|---|
| Cariou, Bertrand 2020 | -0.0513 | 0.1206 | 51.7% | 0.95 [0.75, 1.20] |
| Chen, Yuchen 2020 | 0.2201 | 0.409 | 4.5% | 1.25 [0.56, 2.78] |
| Cheng, Xu 2020 | 0.5188 | 0.952 | 0.8% | 1.68 [0.26, 10.86] |
| Cheng, Xu 2020 | -0.1625 | 0.1693 | 26.2% | 0.85 [0.61, 1.18] |
| Do, Jun Young 2020 | -0.0834 | 0.2533 | 11.7% | 0.92 [0.56, 1.51] |
| Gao, Yongchao 2020 | 1.3773 | 0.6856 | 1.6% | 3.96 [1.03, 15.20] |
| Kim, Mi Kyung 2020 | -0.734 | 0.4728 | 3.4% | 0.48 [0.19, 1.21] |
| **Total (95% CI)** | | | **100.0%** | **0.93 [0.79, 1.11]** |

Heterogeneity: Chi² = 7.64, df = 6 (P = 0.27); I² = 21%
Test for overall effect: Z = 0.78 (P = 0.44)

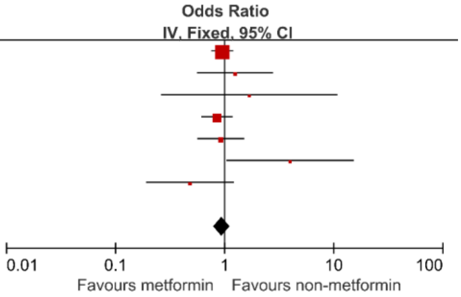


Figure S7. Forest plot of the association between metformin and poor composite outcomes of COVID-19 for diabetic patients

| Study or Subgroup | log[Odds Ratio] | SE | Weight | Odds Ratio IV, Random, 95% CI |
|---|---|---|---|---|
| Cariou, Bertrand 2020 | -0.1625 | 0.2246 | 34.3% | 0.85 [0.55, 1.32] |
| Fadini, Gian Paolo 2020 | -0.2549 | 1.1141 | 11.7% | 0.77 [0.09, 6.88] |
| Kim, Mi Kyung 2020 | 0.3853 | 0.6016 | 22.8% | 1.47 [0.45, 4.78] |
| Solerte, Sebastiano Bruno 2020 | -1.4697 | 0.3319 | 31.2% | 0.23 [0.12, 0.44] |
| **Total (95% CI)** | | | **100.0%** | **0.63 [0.26, 1.56]** |

Heterogeneity: Tau² = 0.57; Chi² = 13.00, df = 3 (P = 0.005); I² = 77%
Test for overall effect: Z = 0.99 (P = 0.32)

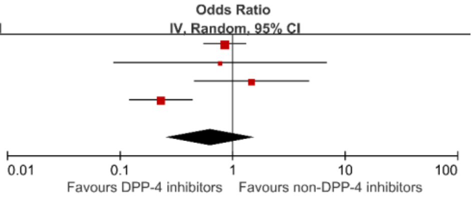


Figure S8. Forest plot of the association between DPP-4i and mortality risk of COVID-19 for diabetic patients

| Study or Subgroup | log[Odds Ratio] | SE | Weight | Odds Ratio IV, Fixed, 95% CI |
|---|---|---|---|---|
| Cariou, Bertrand 2020 | 0.01 | 0.1519 | 80.2% | 1.01 [0.75, 1.36] |
| Fadini, Gian Paolo 2020 | 0.7453 | 0.7671 | 3.1% | 2.11 [0.47, 9.48] |
| Kim, Mi Kyung 2020 | 0.0488 | 0.4438 | 9.4% | 1.05 [0.44, 2.51] |
| Rhee, Sang Youl 2020 | -1.0161 | 0.5033 | 7.3% | 0.36 [0.13, 0.97] |
| **Total (95% CI)** | | | **100.0%** | **0.96 [0.74, 1.26]** |

Heterogeneity: Chi² = 4.96, df = 3 (P = 0.17); I² = 39%
Test for overall effect: Z = 0.28 (P = 0.78)

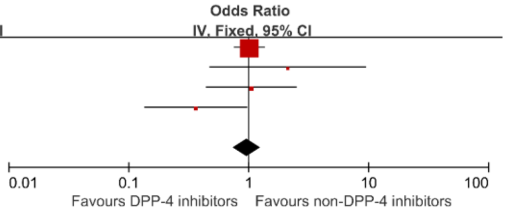


Figure S9. Forest plot of the association between DPP-4i and poor composite outcomes of COVID-19 for diabetic patients

Table S1. search terms and strategies

| | |
|---|---|
| Metformin | Metformin OR Dimethylbiguanidine OR Dimethylguanylguanidine OR Glucophage |
| sulfonylurea | Gliclazide or Gliklazid or Diamicron or Diaglyk or Glyade or Diaikron or Diabrezide or Glimepiride or glimepiride or Amaryl or Amarel or Gliquidone or glikvidon or glycvidon or Glurenorm or Beglynor or Beglynora or Glurenor or Glipizide or Glyburide or Glypidizine or Glidiazinamide or Glydiazinamide or Glucotrol or Minidiab or Mindiab or Minodiab or Melizide or Ozidia or Glupitel |
| Thiazolidinediones | TZDs OR Thiazolidinediones OR Rosiglitazone OR Rosiglitazone Maleate OR Avandia OR Pioglitazone OR Pioglitazone Hydrochloride OR Actos |
| Glinides | Glinides OR Repaglinide OR repa-glinide OR NovoNorm OR GlucoNorm OR Prandin OR Nateglinide OR Senaglinide OR Nate-Glinide OR Fastic OR Starsis OR Starlix OR Mitiglinide OR miti-glinide |
| a-glycosidase inhibitors | a-glycosidase inhibitors OR Acarbose OR Glumida OR Glucobay OR Glucor OR Prandase OR Precose Voglibose OR Basen Miglitol OR Diastabol OR Plumarol OR Glyset |
| DPP-4 Inhibitors | Dipeptidyl-Peptidase IV Inhibitors OR DPP-4 Inhibitors OR Sitagliptin OR Januvia OR Saxagliptin OR Onglyza OR Vildagliptin OR Galvus OR Alogliptin OR nesina OR Tiglitatin OR Linagliptin OR Tradjenta OR Trajenta |
| GLP-1 Receptor Agonists | Glucagon-Like Peptide-1 Receptor Agonists OR GLP-1 Receptor Agonists OR Exenatide OR Bydureon OR Exendin-4 OR Byetta OR Liraglutide OR Victoza OR Saxenda |
| SGLT-2 inhibitors | Sodium-glucose co-transporter 2 inhibitors OR SGLT-2 inhibitors OR Dapagliflozin OR farxiga OR forxiga OR Empagliflozin OR Jardiance OR Canagliflozin OR Invokana |
| hypoglycemic agents | antidiabetic drugs OR hypoglycemic agents OR hypoglycemic drug |
| COVID-19 | COVID-19 OR COVID19 OR Coronavirus Disease 2019 OR 2019 novel coronavirus OR 2019-nCoV OR SARS-COV-2 |

Table S2. effects of metformin vs. non-metformin: composition of poor outcomes

| Study | ARDS | DIC | ICU | ECMO | death | tracheal intubation | progress | organ dysfunctions | sepsis | septic shock |
|---|---|---|---|---|---|---|---|---|---|---|
| Cariou, Bertrand 2020 | | | | | √ | √ | | | | |
| Chen, Yuchen 2020 | | | | | √ | | √ | | | |
| Cheng, Xu 2020 | √ | √ | | | | | | | | |
| Do, Jun Young 2020 | | | | √ | | | | √ | | |
| Gao, Yongchao 2020 | √ | | √ | | | | | √ | √ | √ |
| Kim, Mi Kyung | | | √ | √ | | √ | | √ | | |
